# Elevated remnant cholesterol improves prognosis of patients with ischemic stroke and malnutrition: a cohort-based study

**DOI:** 10.1101/2024.07.31.24311324

**Authors:** Huicong Niu, Yong Wang, Ning Yang, Min Chu, Xueyu Mao, Daosheng Wang, Jing Zhao

## Abstract

**Background:** Aggressive lipid-lowering therapy is recommended for patients with ischemic stroke; however, lipid paradox has been reported in several clinical studies. The mechanism of lipid paradox remains uncertain, and nutrition maybe one explanation. In this prospective cohort study, we explored the associations between baseline remnant cholesterol (RC) concentrations and clinical outcomes in patients with ischemic stroke, stratified by nutritional status.

**Materials and Methods:** A total of 5257 patients with ischemic stroke were recruited for analysis. The Controlling Nutritional Status (CONUT) score was utilized to investigate the risk of malnutrition. Individuals were classified into 4 groups based on their CONUT score. Poor outcomes and all-cause mortality were compared among patients with varied nutritional status and RC levels.

**Results:** Patients with moderate-severe malnutrition had the highest incidences of in-hospital complications, including pulmonary infection, renal dysfunction, and hemorrhagic transformation, and the highest rates of poor outcomes (61.3%, *P*<0.001) and all-cause mortality (32.8%, *P* < 0.001) during the 3-month follow-up period. Baseline higher RC level was an independent protective factor of adverse clinical outcomes for patients with any degree of malnutrition, which was not observed in patients without malnutrition. In addition, compared with the moderate-severe malnourished with RC <0.471 mmol/L, the adjusted ORs for poor outcomes and all-cause mortality were 0.805 (0.450–1.438) and 0.898 (0.502-1.607) for participants with 0.471-0.632 mmol/L, 0.259 (0.095-0.704) and 0.222 (0.061-0.810) for 0.633-0.868 mmol/L, and 0.160 (0.037–0.689) and 0.202 (0.042-0.967) for ≥ 0.869 mmol/L, prospectively.

**Conclusion:** Lipid paradox was only observed in the malnourished patients with ischemic stroke. Strict lipid reduction therapy is still recommended for patients with ischemic stroke and good nutritional status. However, when treating patients at any risk of malnutrition, the improvement of nutritional status may be more crucial than aggressive lipid control.

## Introduction

Lipid disorders, especially those involving elevated cholesterol and low-density lipoprotein cholesterol (LDL-C), are major causal risk factors in the onset and progression of atherosclerotic cerebrovascular disease^1^. Especially, the atherogenic potential of remnant cholesterol (RC), carried in chylomicrons, chylomicron remnants, very low-density lipoprotein (VLDL), and intermediate-density lipoprotein (IDL), is being increasingly acknowledged^2–5^. Higher RC levels may partially explain the significant residual risk factors for atherosclerosis among statin-treated individuals, even in the context of aggressive lipid-lowering treatment to reach LDL-C targets^6, 7^. For example, the Copenhagen General Population Study has adequately evidenced that individuals with high RC concentrations have a higher risk of ischemic stroke, and indicated that, for effectively preventing ischemic stroke, randomized clinical trials with RC-lowering therapy in individuals with high concentrations are desperately needed^8^.

Although an essential risk factor, the lipid profile was brought again into the spotlight in patients with cardio-cerebrovascular diseases as the result of recent studies manifesting a phenomenon called “lipid paradox”, meaning a paradoxical relationship between the lipidic profile and functional outcome of patients^9–11^. Several hypotheses were stated regarding the physiopathological mechanism responsible for this paradox, including the increased rupture resistance of the vessels, the maintenance of vascular integrity, neuroplasticity’s role in developing new synapses, and neuroprotective effects in the central nervous system^1, 9^. However, few studies have attributed the lipid paradox to the presence of malnutrition, and no existing research has been made to clarify lipid paradox utilizing RC, rather than LDL-C.

In this single-center prospective cohort study, we observed the relationship between baseline RC levels and clinical outcomes in hospitalized patients with ischemic stroke, stratified by nutritional status. We applied the Controlling Nutritional Status (CONUT) score to access the risks of malnutrition, and elucidate the association among nutritional status, RC levels, and clinical outcomes in patients with ischemic stroke.

## Materials and Methods

### Study participants

The participants were selected from the Minhang Stroke Cohort study, which was established based on patients with ischemic stroke at Minhang Hospital in Shanghai, China. Briefly, from January 2018 to December 2022, 7323 consecutive patients aged 18 years or older, diagnosed as ischemic stroke, were prospectively invited from the department of neurology at Minhang Hospital, and patients with time from onset to admission exceeding 7 days or diagnosed as tumors were excluded. Finally, a total of 6892 patients were enrolled in the Minhang Stroke Cohort. For the present study, additional exclusion criteria were as follows: prestroke modified Rankin Scale (mRS) >2 (n=260); acute infection within 2 weeks before admission (n=464); lack measurements of lymphocyte count, albumin, total cholesterol (TC), triglyceride (TG), HDL-C, and LDL-C (n=470); or loss to follow-up (n=441). In all, 5257 individuals were included in the analysis as shown in the flow diagram (**Fig. S1**). The study was approved by the Institutional Ethics Committee of Minhang Hospital of Fudan University and complied with the Declaration of Helsinki.

### Baseline data collection

The baseline data of participants who were enrolled in the Minhang Stroke Cohort study was obtained by well-trained research coordinators through direct interviews or medical records. The collected information included demographics, medical history, clinical characteristics, and lifestyle. Stroke severities were evaluated with the National Institutes of Health Stroke Scale (NIHSS) score by experienced neurologists at admission, and mRS score was applied to assess patients’ functional independence^12^. The ischemic stroke subtype was identified as large-artery atherosclerosis (LAA), cardioembolism (CE), small-vessel occlusion (SAO), stroke of other determined etiology, and stroke of undetermined etiology based on the Trial of ORG 10172 in Acute Stroke Treatment (TOAST) criteria^13^.

### Laboratory analyses

Fasting blood samples of the patients were timely collected within 24 hours of admission, and TC, TG, high-density lipoprotein cholesterol (HDL-C), glucose, albumin, estimated glomerular filtration rate (eGFR), and blood cells were measured with a Cobas 8000 automatic analyzer (Roche Diagnostics, Indianapolis, Indiana) by laboratory personnel unaware of the clinical data. LDL-C was calculated using the Friedewald equation^14^, unless TG were >4.5 mmol/L (400 mg/dL), where it was measured directly. The formula RC=TC–LDL-C–HDL-C was used to calculated the RC values^15^. This means that in individuals with TG≤4.5 mmol/ L (400 mg/dL), RC was TG (in mmol/L) divided by 2.2, as calculated by the Friedewald equation, while at TG>4.5 mmol/L RC was TC minus measured LDL-C minus measured HDL-C. Nutritional status was graded by the CONUT score, which was computed with the following formula: CONUT score=albumin score+total cholesterol score+lymphocyte count score, and scores of 0 to 1, 2 to 4, 5 to 8, 9 to 12 were defined as normal, mild, moderate, and severe malnutrition status, respectively^16^ (**Table S1**). To examine the prognostic impact of the RC level on patients with different risks of malnutrition, we stratified the study cohort by serum RC median into high RC group (>0.632 mmol/L) and low RC group (≤0.632 mmol/L).

### Endpoints

The participants recruited in the study were followed up at 3 months after ischemic stroke by trained neurologists through telephone or face-to-face interviews. Information on dead was acquired from their relatives, which was furtherly verified by the death certificate from the local citizen registry or the attended hospital. Mortality included death from all cause, and the definition of poor outcomes was mRS score 3-6. All study outcomes were evaluated and judged by a committee who was blind to the clinical characteristics of the participants.

### Statistical analyses

The study population were stratified into three categories base on their nutritional status: without malnutrition, mild malnutrition, and moderate-severe malnutrition. Continuous data were expressed as mean ± standard deviation, and categorical variables are summarized as percentages. Binary and continuous variables were compared using χ^2^ test and one-way ANOVA. We utilized multivariate logistic regression analyses to explore whether the high RC level (>0.632 mmol/L) was associated with better clinical outcomes of ischemic stroke. The covariates included in the multivariable models were age, gender, admission NIHSS scores, reperfusion therapy, smoking history, drinking history, hypertension, diabetes mellitus, prior stroke, hyperlipidemia, coronary artery disease, atrial fibrillation, stroke subtype, and use of statin.

Restricted cubic splines (RCSs) with 4 knots (5th, 35th, 65th, and 95th percentiles) of serum RC were carried out to evaluate the non-linearity association between serum RC on a continuous scale and prognosis of ischemic stroke. In addition, we divided the population into quartiles based on their RC concentrations to investigate the association between serum RC quartiles and clinical outcomes of patients in different risks of malnutrition with multivariate logistic regression analyses. Subgroup analyses were also utilized to investigate the modifiable effect of certain stroke risk factors (age, gender, admission NIHSS scores, reperfusion therapy, smoking history, drinking history, hypertension, diabetes mellitus, prior stroke, hyperlipidemia, coronary artery disease, atrial fibrillation, stroke subtype, and use of statin) on the association between serum RC and clinical outcomes of ischemic stroke. Interactions between serum RC and subgroup variables on poor outcomes and all-cause mortality were estimated in the models with interaction terms using the likelihood ratio test, adjusting for the aforementioned covariates. All *P* values were two-tailed, and a significant level of 0.05 was used.

## Results

### Baseline characteristics

Most baseline characteristics were well-balanced between patients invited in the present study and all participants of the Minhang Stroke Cohort study, hinting that those recruited were principally representative of the total patients form the Minhang Stroke Cohort (**Table S2**). As shown in **Table 1**, Among 5257 participants (3398 male and 1859 female) summarized by different nutritional status, those with moderate-severe malnutrition tended to be older, nonsmokers, and nondrinkers; to have higher prevalence of history of coronary artery disease and atrial fibrillation; to have higher admission NIHSS score; and to have higher prevalence of statin usage. However, they were also likely to have lower diastolic pressure and a lower prevalence of history of hyperlipidemia. The hemoglobin level and baseline eGFR significantly decreased in patients with moderate-severe malnutrition. Nutritional parameters, including albumin, lymphocyte count, TC, TG, and LDL-C levels, differed significantly in various nutritional status. The group without malnutrition included more patients with a baseline RC > 0.632 mmol/L.

**Table 1.**
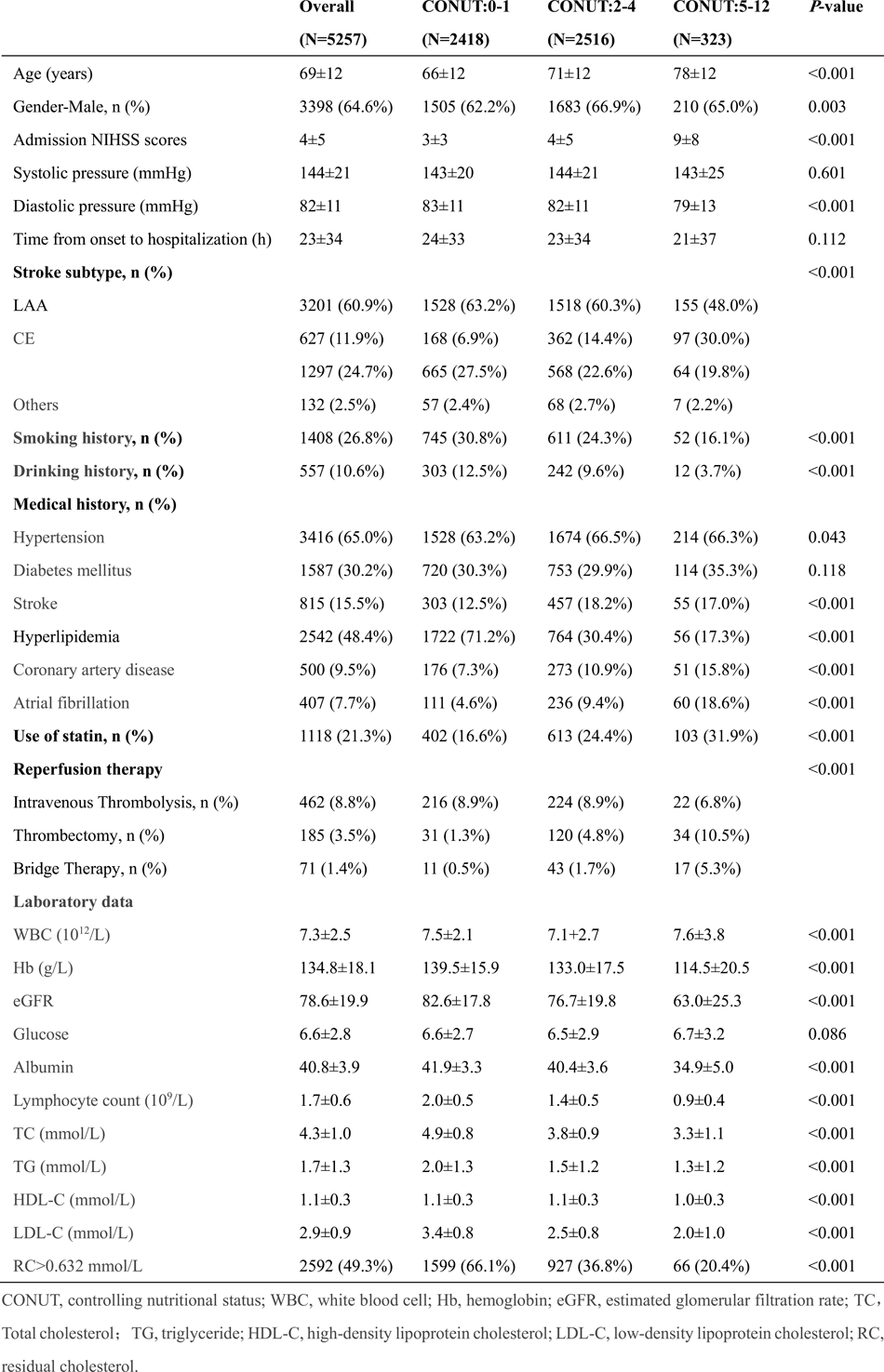
Baseline characteristics of the Study Population based on nutritional status.

### Clinical outcomes stratified by nutritional status and RC levels

The highest prevalence of in-hospital complications, including pulmonary infection (23.5%), renal dysfunction (13.0%), and hemorrhagic transformation (5.0%), were found in patients with moderate-severe malnutrition (all *P*<0.001; **Table 2**). There were also longer hospital length of stay (11±9 days) and total hospitalization cost (40,327±53,465 Chinese Yuan Renminbi (RMB)) in the moderate-severe malnourished (all *P*<0.001; **Table 2**). At 3 months after ischemic onset, a total of 1644 (31.3%) participants experienced poor outcomes and 423 (8.0%) patients died. Malnutrition severity was significantly associated with an increased incidence of poor outcomes (mild malnutrition: 34.5%, moderate-severe malnutrition: 61.3%, *P*<0.001) and 3-month mortality (mild malnutrition: 8.3%, moderate-severe malnutrition: 32.8%, *P*<0.001) for hospitalized patients with ischemic stroke (**Table 2 and Fig. S2**).

**Table 2.**
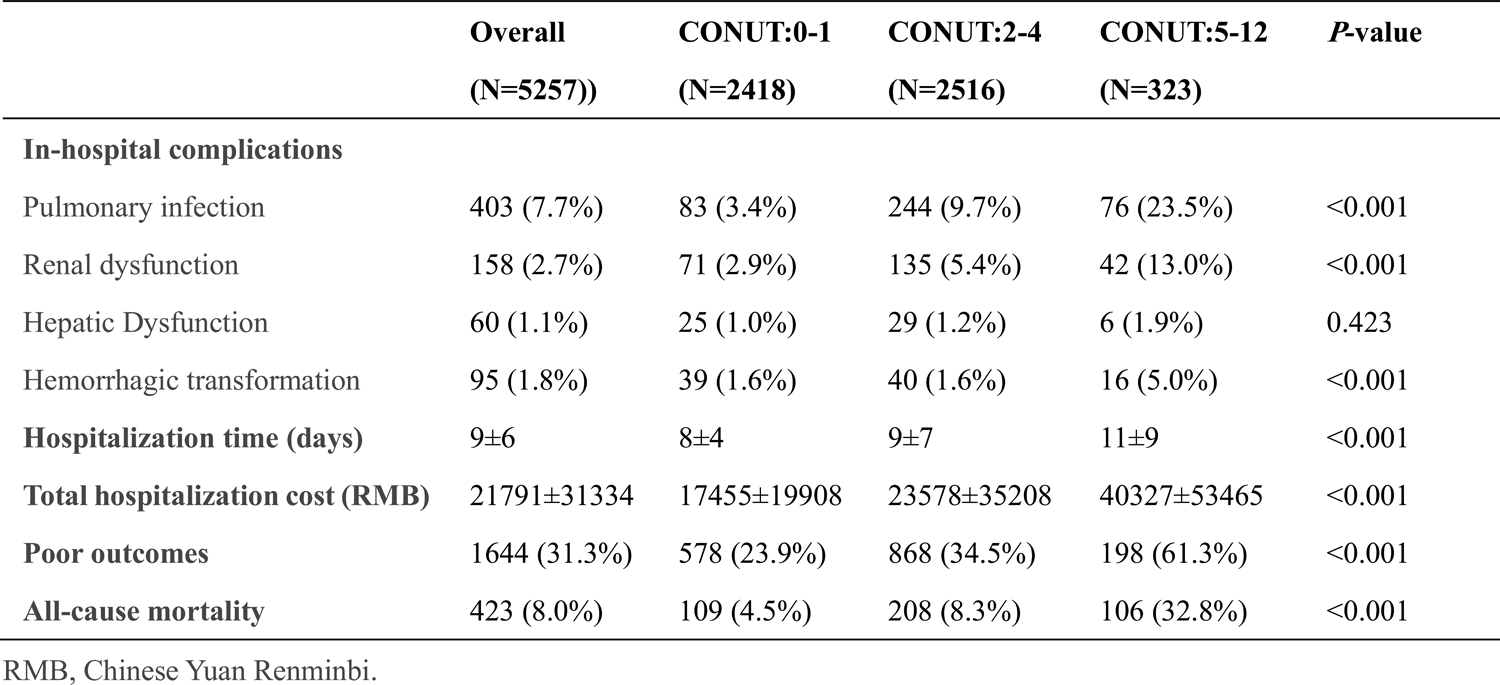
Outcomes according to nutritional status.

The clinical outcomes of patients, stratified by nutritional status and RC level (0.632 mmol/L as the median), were revealed in **Fig. 1**. The overall study population (**Fig. 1a**) and the participants with any degree of malnutrition (**Fig. 1c** and **d**) displayed significantly better outcomes and lower all-cause mortality when baseline RC levels exceeded 0.632 mmol/L, but the protective effect of high RC level was not observed in patients without malnutrition (**Fig. 1b**).

**Fig. 1.**
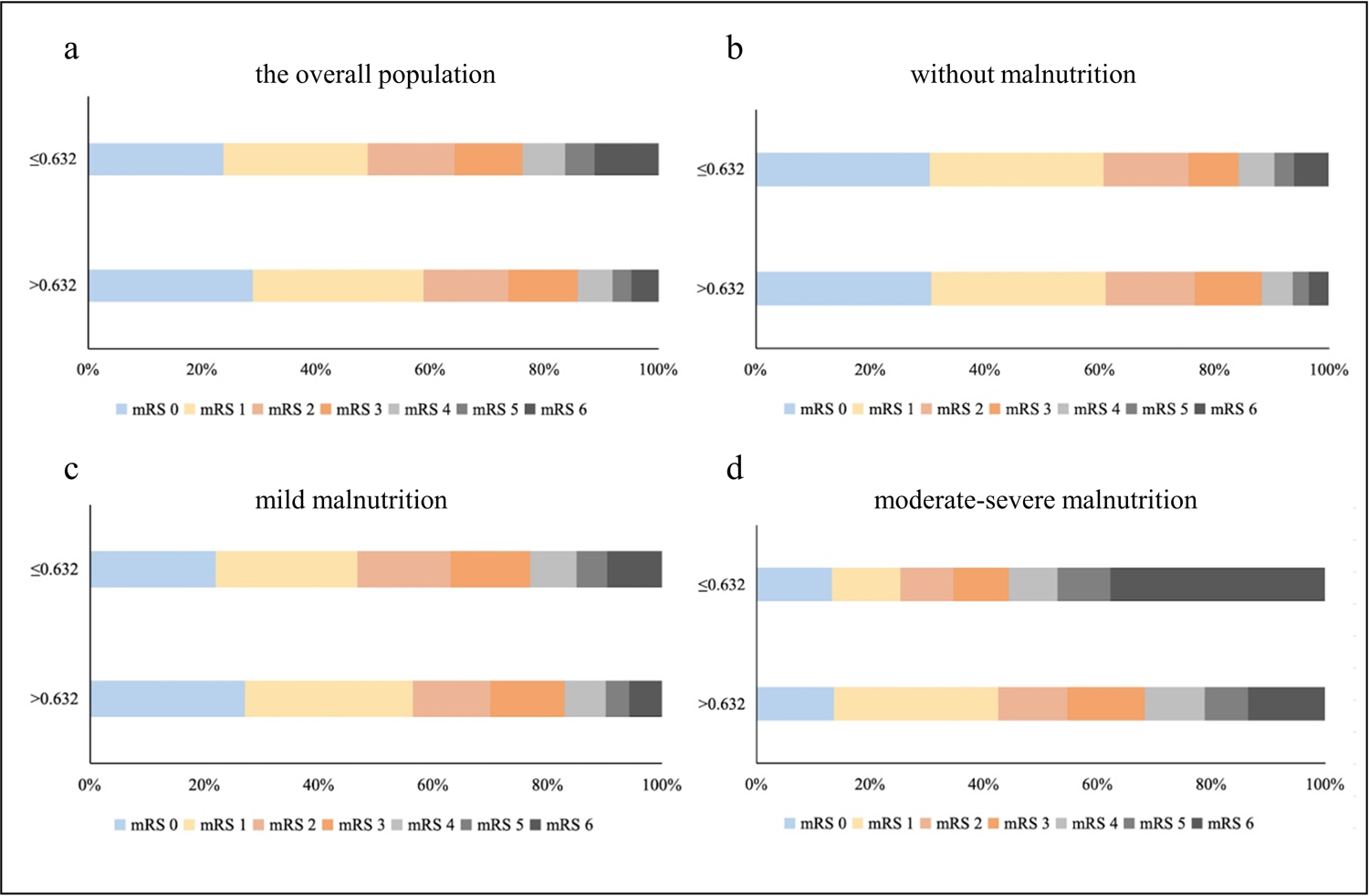
The distribution of stroke outcomes in different population stratified by nutritional status and RC level: the overall population (a), without malnutrition (b), with mild malnutrition (c), and moderate-severe malnutrition (d). mRS, modified Rankin Scale.

The adjusted odds ratio (OR) according to the median of serum RC levels were shown in **Table 3 and 4**. For poor outcomes and all-cause mortality in the overall population, the multivariable adjusted ORs for patients with baseline RC> 0.632 mmol/L versus the group with RC≤0.632 level were 0.789 (0.678-0.919) and 0.547 (0.417-0.718), prospectively (all *P*<0.05). Similar results were observed in patients with mild malnutrition and moderate-severe malnutrition, and the worse the nutritional status, the higher the OR. In contrast, patients without malnutrition showed a nonsignificant difference in adverse clinical outcomes (0.971 (0.766-1.230) for poor outcomes and 0.723 (0.454-1.151) for all-cause mortality, regardless of RC level.

**Table 3.**
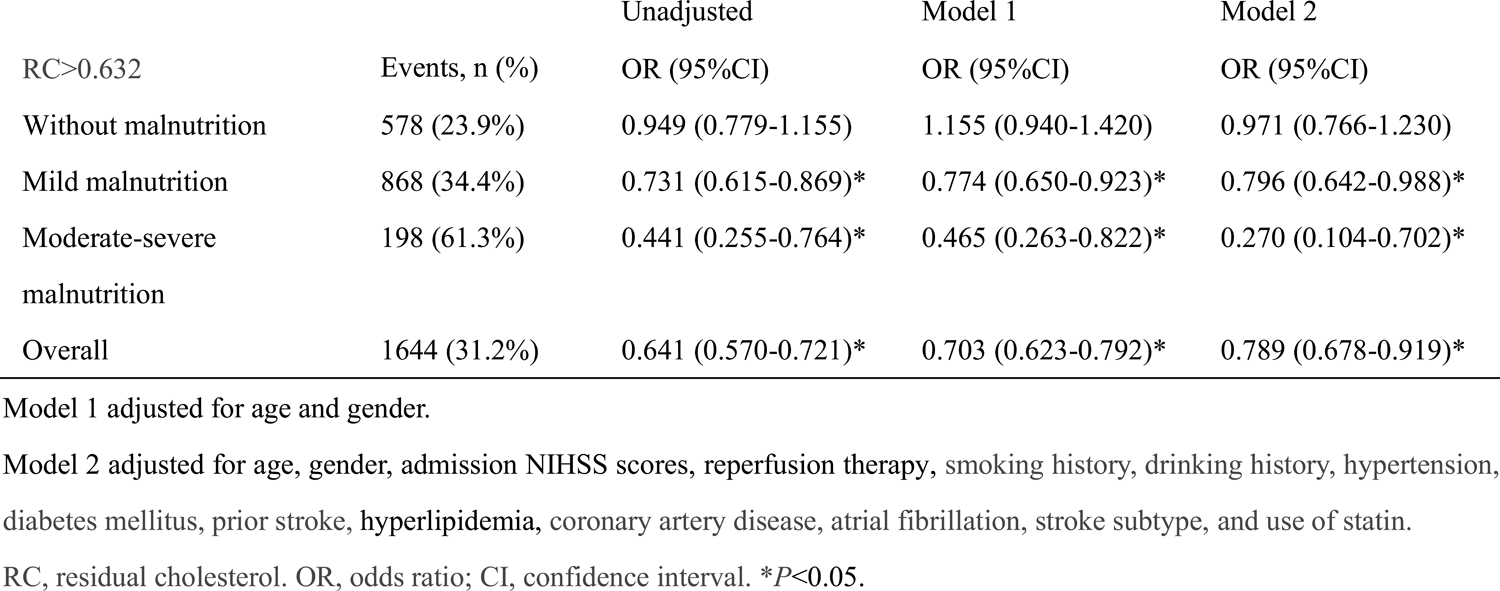
Multivariable logistic regression results for poor outcomes in patients with ischemic stroke (whole study cohort, n=5257)

**Table 4.**
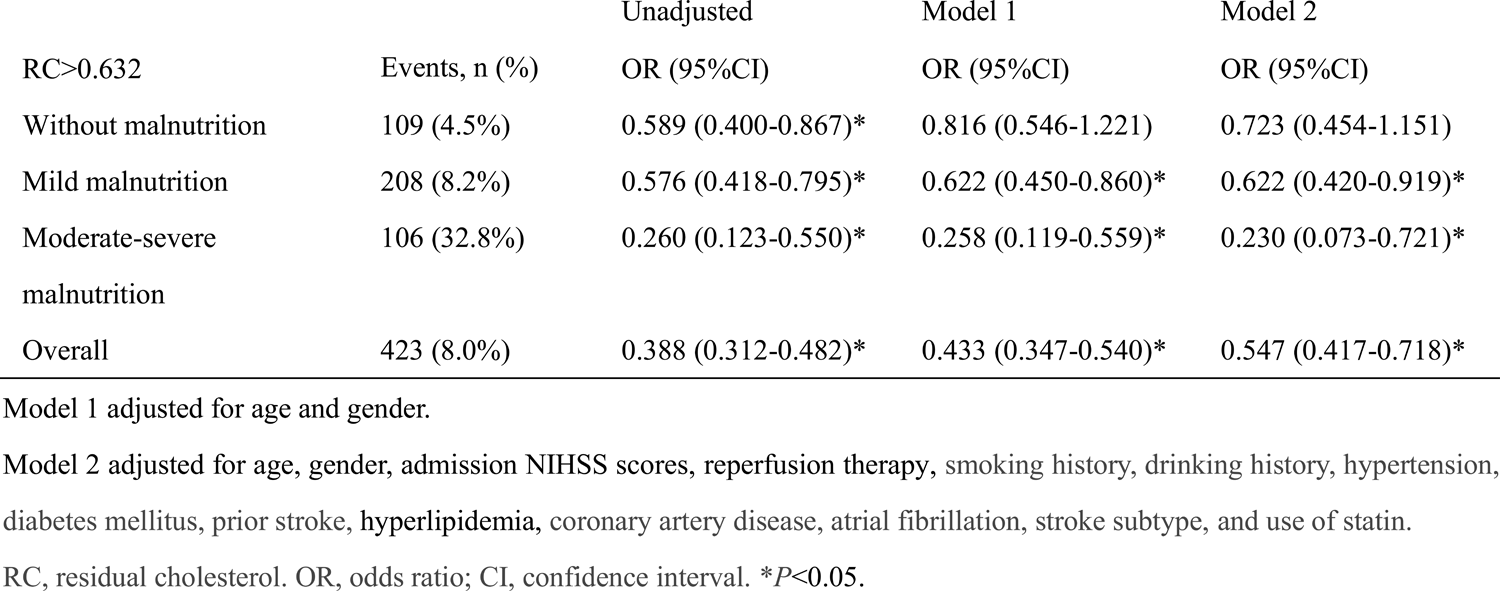
Multivariable logistic regression results for all-cause mortality in patients with ischemic stroke (whole study cohort, n=5257)

Serum RC and adverse outcome risk of ischemic stroke in the overall population Actually, lipid paradox existed the individuals with ischemic stroke. As shown by the RCSs (**Fig. 2**), higher concentrations of RC were significantly associated with decreased risk of poor outcomes and during the follow-up of 3 months in the overall population. This association showed a trend of increasing risk that eventually leveled off when remnant cholesterol level exceeded 1.1 mmol/L. The adjusted ORs for poor outcomes and all-cause mortality were 0.815 (0.690–0.962) and 0.807 (0.628-1.038) for participants with 0.471-0.632 mmol/L, 0.732 (0.608–0.880) and 0.494 (0.358-0.682) for 0.633-0.868 mmol/L, and 0.604 (0.482–0.575) and 0.411 (0.274-0.616) for ≥ 0.869 mmol/L, compared with individuals with RC<0.471 mmol/L, prospectively (**Fig. 3**).

**Fig. 2.**
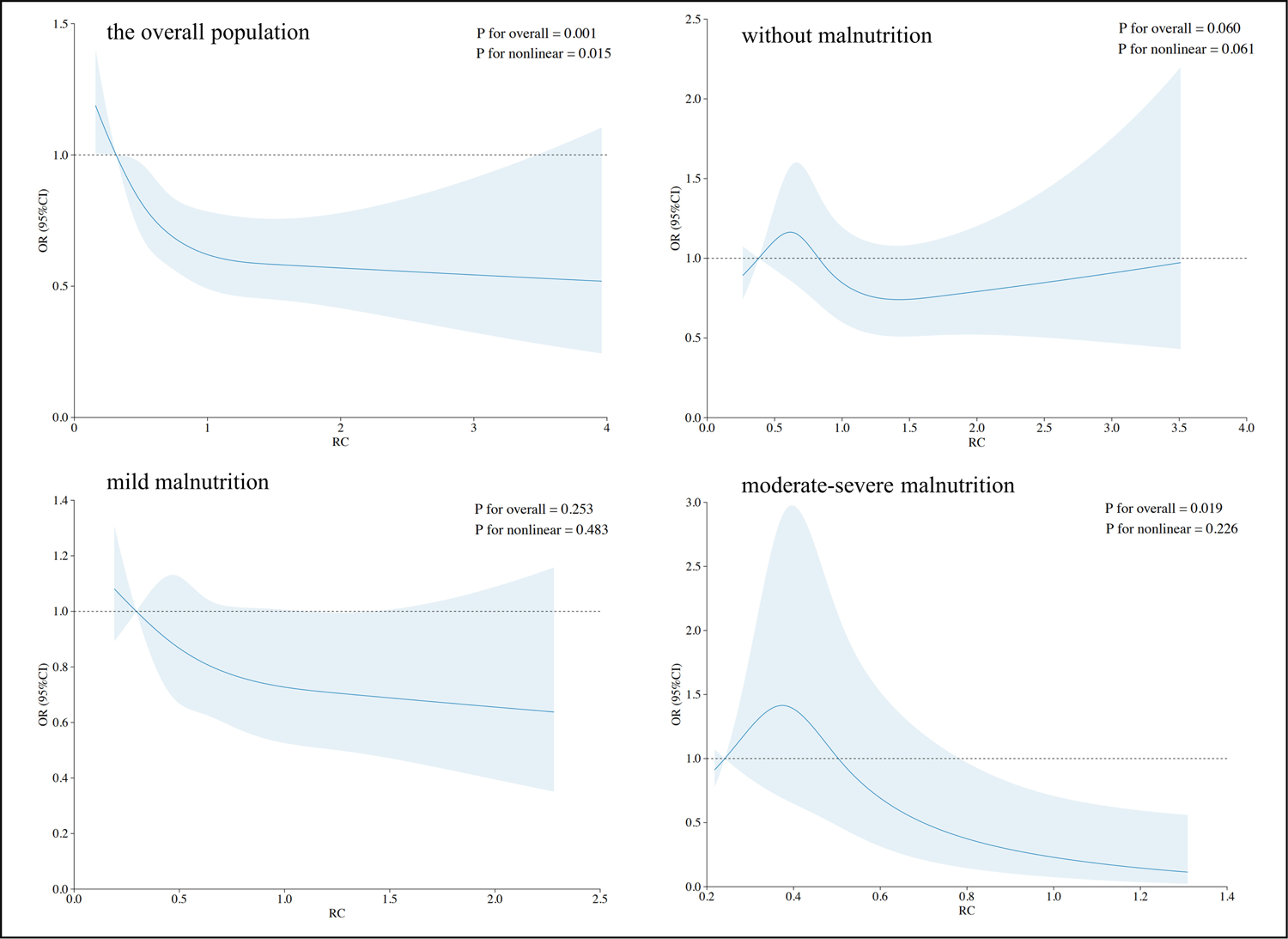
Distribution of RC level and its association between RC with poor outcomes. Restricted cubic splines of RC with 4 predefined knots (5th, 35th, 65th, and 95th percentiles) to evaluate the non-linearity association of RC on a continuous scale and risk of poor outcomes. These models were adjusted for age, gender, admission NIHSS scores, reperfusion therapy, smoking history, drinking history, hypertension, diabetes mellitus, prior stroke, hyperlipidemia, coronary artery disease, atrial fibrillation, stroke subtype, use of statin. RC, residual cholesterol.

**Fig. 3.**
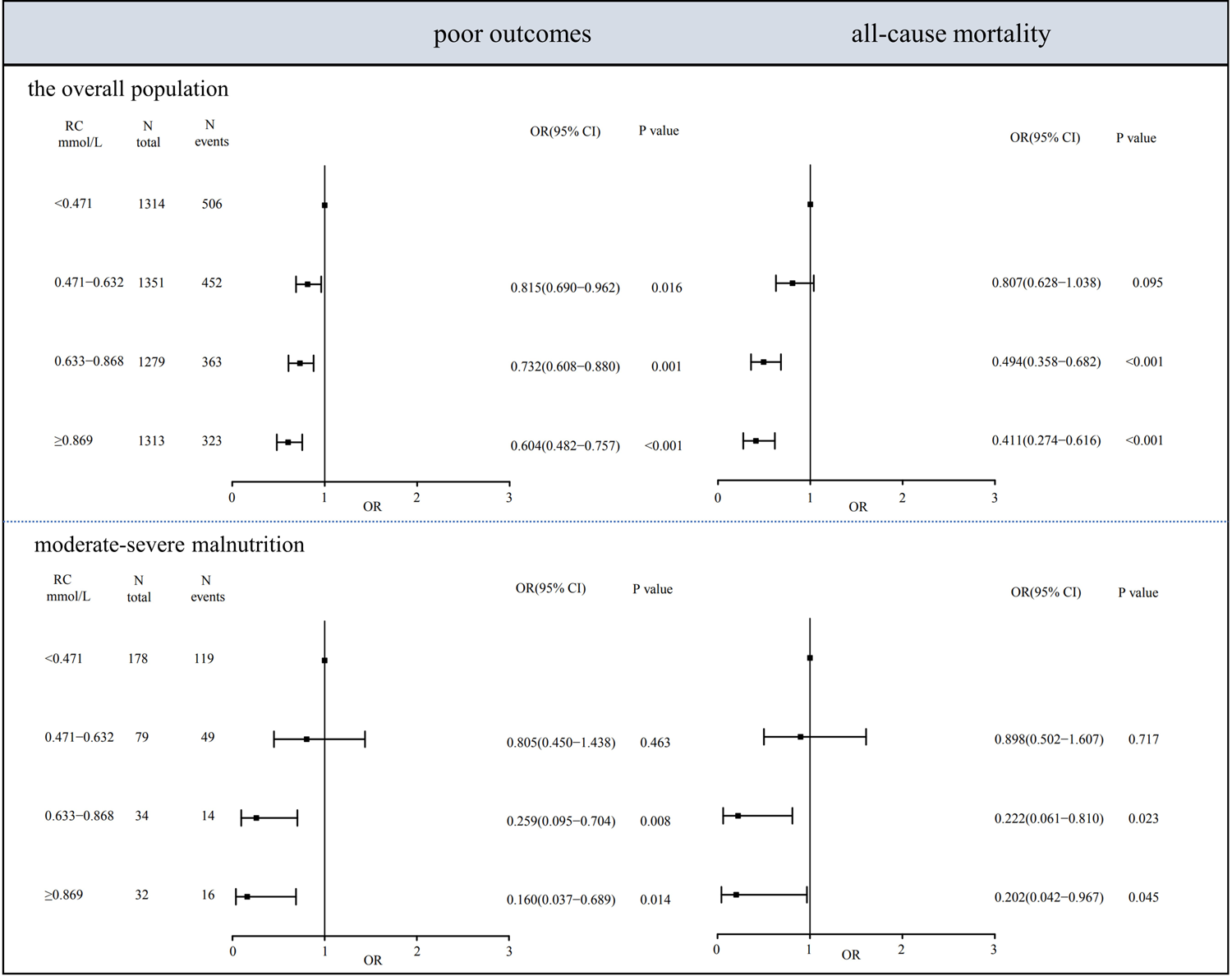
Association between RC in quartiles and risk of poor outcomes/all-cause mortality in the overall population and the moderate-severe malnourished. Adjusted for age, gender, admission NIHSS scores, reperfusion therapy, smoking history, drinking history, hypertension, diabetes mellitus, prior stroke, hyperlipidemia, coronary artery disease, atrial fibrillation, stroke subtype, use of statin. RC, residual cholesterol. OR, odds ratio; CI, confidence interval.

Serum RC and adverse outcome risk of ischemic stroke in patients without malnutrition The poor outcomes (*P*=0.060) and all-cause mortality (*P*=0.120) did not differ significantly between patients in good nutritional status with different baseline RC concentrations, which was revealed in **Fig. 2 and S3**. The adjusted ORs for poor outcomes and all-cause mortality were 1.051 (0.740–1.493) and 1.278 (0.670-2.435) and for participants with 0.471-0.632 mmol/L, 1.163 (0.823-1.643) and 0.908 (0.459-1.796) for 0.633-0.868 mmol/L, and 0.785 (0.544–1.333) and 0.732 (0.350-1.533) for ≥0.869 mmol/L, compared with participants with RC <0.471 mmol/L, prospectively (**Fig. S4**).

Serum RC and adverse outcome risk of ischemic stroke in patients with malnutrition For the mild malnourished, although nonsignificant protective effect was found in the RCSs (**Fig. 2 and S3**), increasing negative association between serum RC level and all-cause mortality risk could be observed. Compared with participants in Quartile 1, ORs for the other three quartiles were 0.935 (0.662–1.320), 0.632 (0.400–0.970), and 0.418 (0.224–0.781) for the individuals with mild malnutrition (**Fig. S4**).

In addition, RC levels < 0.3 mmol/L showed hazardous effect in individuals with moderate-severe malnutrition, which turned into a protective effect when RC levels ≥ 0.5 mmol/L (**Fig. 2**). Furthermore, adverse outcomes were progressively higher in the moderate-severe malnourished with lower RC levels. The adjusted ORs for poor outcomes and all-cause mortality were 0.805 (0.450–1.438) and 0.898 (0.502-1.607) and for participants with 0.471-0.632 mmol/L, 0.259 (0.095-0.704) and 0.222 (0.061-0.810) for 0.633-0.868 mmol/L, and 0.160 (0.037–0.689) and 0.202 (0.042-0.967) for ≥ 0.869 mmol/L, compared with participants with RC <0.471 mmol/L, prospectively (**Fig. 3**).

### Subgroup and sensitivity analyses

Besides, subgroup analyses were conducted to investigate the potential modification of several stroke risk factors, including age, gender, admission NIHSS scores, reperfusion therapy, smoking history, drinking history, hypertension, diabetes mellitus, prior stroke, hyperlipidemia, coronary artery disease, atrial fibrillation, stroke subtype, and use of statin, on the association between serum RC levels and the adverse outcomes of ischemic stroke. We demonstrated that a high serum RC level significantly increased the risk of poor outcomes and all-cause mortality in the overall population and subgroups of patients with malnutrition, but not in the nourished. These results were consistent across different clinical subgroups in the study population stratified by nutritional status (**Fig. 4 and S5**).

**Fig. 4.**
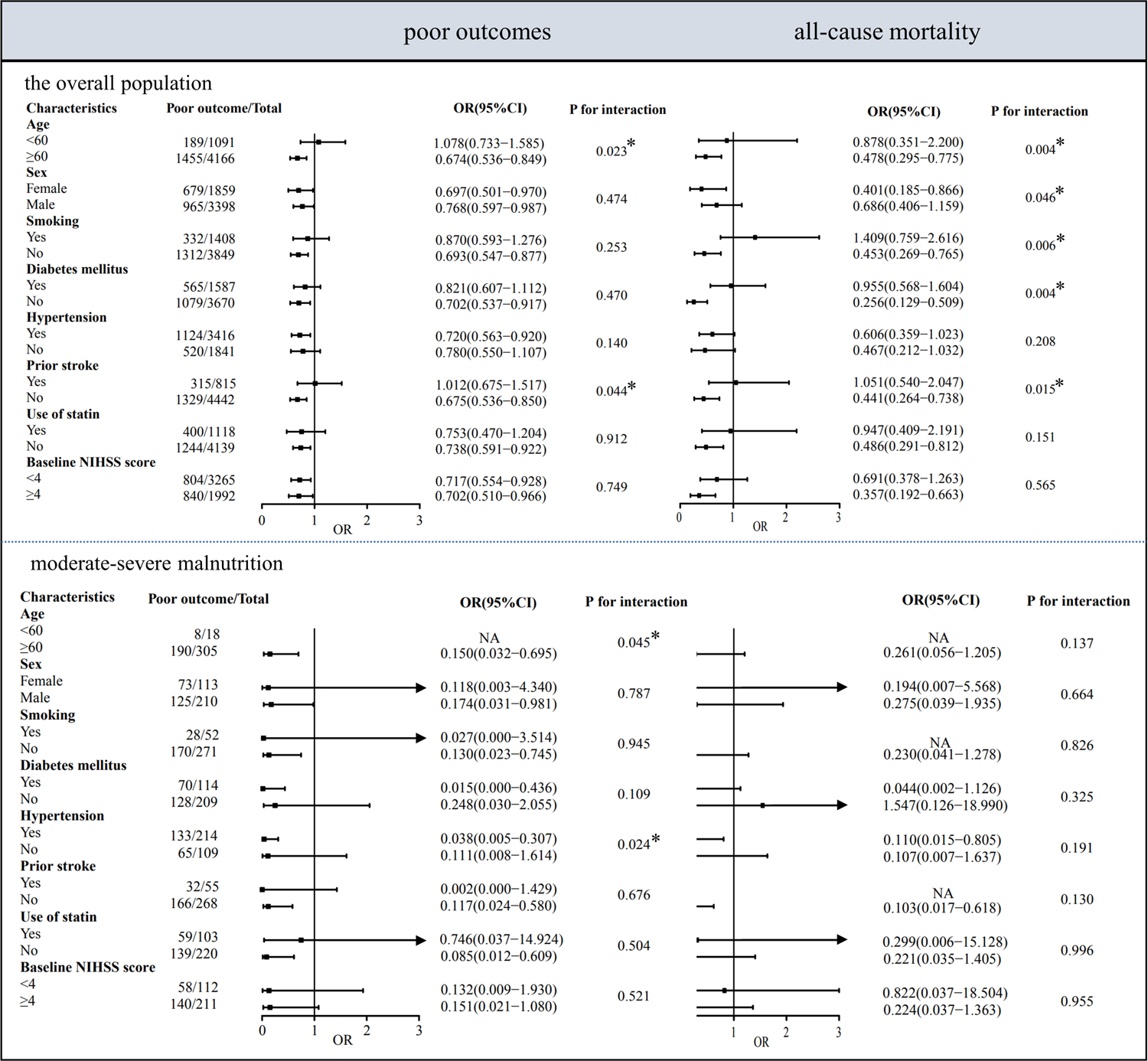
Subgroup analyses of the association between RC and poor outcomes/all-cause mortality in the overall population and the moderate-severe malnourished (quartile 4 vs quartile 1, data for quartiles 2 and 3 not shown). Adjusted for age, gender, admission NIHSS scores, reperfusion therapy, smoking history, drinking history, hypertension, diabetes mellitus, prior stroke, hyperlipidemia, coronary artery disease, atrial fibrillation, stroke subtype, use of statin, unless the variable was used as a subgroup variable, RC, residual cholesterol. OR, odds ratio; CI, confidence interval.

## Discussion

In this prospective cohort study of 5257 cases of ischemic stroke, patients with malnutrition presented a higher incidence of in-hospital complications, including pulmonary infection, renal dysfunction, and hemorrhagic transformation. Higher RC level was an independent protective factor for the clinical outcomes of patients with ischemic stroke. Of note, the lipid paradox was only seen in the malnourished individuals with ischemic stroke. Baseline higher RC level was associated with decreased incidence of all-cause mortality in the mild malnourished individuals. Moreover, baseline higher RC level was an independent protective factor for both poor outcomes and all-cause mortality in the population with moderate-severe malnutrition, but not in the group without malnutrition.

Stroke is the foremost cause of adult disability and a major cause of mortality around the world, and ischemic stroke has been considered an important public health issue that threatens people’s health and well-being^17^. Hence, it is vital to prevent the disease if possible, with the most important being smoking cessation, high blood pressure, treatment of atrial fibrillation or hyperlipidemia. Besides LDL-C, there is increasing evidence that RC is also associated causally with atherosclerotic plague^18–20^. RC is the main TG-rich lipoproteins, which can be easier metabolized and might mechanistically penetrate the arterial wall, where they get trapped and lead to atherosclerosis^8, 21^. Observational and causal (genetic) population-based studies have found associations between increased RC level and elevated risk of myocardial infarction and ischemic heart disease^22, 23^, while observational studies have shown that increased RC was also associated with elevated risk of ischemic stroke^8^. Furthermore, results from the randomized, controlled REDUCE-IT trial published in 2019 exhibited that when patients with increased TG levels were given 4g of a purified omega-3 fatty acid, icosapent ethyl, major cardiovascular events were reduced by 25% and strokes by 28%^24^.

Although the concept of “the lower the cholesterol, the better the outcome” stands as a well-known slogan for lipid-lowering therapy in patients with ischemic stroke^25^, Several studies have shown that a higher cholesterol level is associated with a better stroke outcome^26–29^. A significant association between elevated admission cholesterol at ischemic stroke onset and improved short-term functional outcome^30^ or 10-year survival^31^ was found in earlier studies. In addition, recent studies revealed the so-called “lipid paradox” also existed in patients with acute coronary syndrome, and the lower LDL-C level on admission predicted a long-term worse prognosis, regardless of statin therapy at discharge^32, 33^. In the present study, we also found that a higher RC level was associated with a decreased incidence of poorer outcomes and all-cause mortality in patients with ischemic stroke (**Table 3, 4 and Fig. 1**).

Despite the uncertainty of mechanism leading to lipid paradox, following explanations have been mentioned. First, cholesterol is a crucial element of intracellular transport, cell membranes, and cell signaling, and lower cholesterol may result in the failure of neuronal cells’ ability to resist acidosis and local hyperosmolarity ischemic stress. Cholesterol is also a precursor of stress hormones, such as cortisol, which contribute to addressing life-threatening stress. Therefore, lower cholesterol might be hazardous under acute injury^9^. In addition, higher serum cholesterol levels were also found to have an anti-inflammatory effect and cholesterol can work as a buffer to neutralize certain free radicals after ischemic attack, resulting in improved tolerance to anoxia^29^. More importantly, the nutrition hypothesis has become a research hotspot in recent years, which holds that the serum cholesterol and TG levels of patients with good nutritional reserve are higher than those of patients with modest nutritional status.^34^. In this work, our results clearly validated that the lipid paradox, utilizing RC as the measurement, was presented only in patients with malnutrition (**Fig. 2, 3, S3, and S4**). In end-stage renal disease, burn injury, and critically ill patients, a change in a single serum protein marker, such as albumin or prealbumin, might represent an acute-phase response and might not accurately reflect the patient’s nutritional status^35–37^. Thus, we evaluated patients’ nutritional status using the COUNT score, which has been demonstrated to be associated with the length of hospitalization, the total hospitalization cost, and risk of complications, and also to predict poor outcome and all-cause mortality in hospitalized patients with ischemic stroke at the 3-month follow-up (**Table 2**). Those results were persistent with the previous studies^38–41^. Moreover, when stratifying patients according to their nutritional status, we found that a higher baseline RC level was an independent protective factor of clinical outcomes among patients with mild malnutrition and moderate-severe malnutrition. The lipid paradox did not exist in patients with good nutritional status. The same phenomenon was also observed in patients with acute myocardial infarction. For example, Ya-Wen Lu et al. performed a single-center retrospective study, recruiting 409 patients with acute myocardial infarction, to explore the relationships between baseline LDL-C concentrations and clinical outcomes according to different nutritional status, and the results showed that lipid paradox was only observed among patients at high-risk of malnutrition^42^. All those results hints that although current clinical practice concentrates on the lipid-lowering therapy and effective control of serum lipids as a crucial approach to preventing or treating ischemic stroke^43, 44^, when treating patients with malnutrition, the improvement of nutritional status may be more beneficial than strict lipid management.

## Study limitation

First, the application of calculated RC may be considered a limitation of our study, resulting from its highly dependent on TG levels, that is, in the main analyses RC was TG in mmol/L divided by 2.2 when TG was ≤4.5 mmol/L (400 mg/dL), while at TG >4.5 mmol/L, RC was total cholesterol minus measured LDL-C minus measured HDL-C. However, calculated and directly measured RC are closely correlated. Therefore, the calculation of RC in this study is considered to be reliable. Second, the present observational study cannot fully eliminate confounding factors, we could not draw conclusions about causality. There could be residual confounding even after extensive adjustment. There is a need for large, randomized trials with RC-lowering therapy in individuals with high RC concentrations according to nutritional status, to see if this can improve the prognosis of patients with ischemic stroke. Third, this research only focused on the baseline RC levels, not the longitudinal lipid changes. The results could not show the influence of individual lipid changes caused by treatment, even though a correction of statin use was done in this work.

## Conclusion

In all, the presence of the lipid paradox differed with varied nutritional status. Baseline high RC level was significantly associated with better outcomes and lower mortality during 3-month follow up in the malnourished patients with ischemic stroke, but not in those with good nutritional status. Lipid-lowering therapy is still suggested for patients with ischemic stroke and good nutrition. Whereas, for patients with malnutrition, especially those with moderate-severe malnutrition, it is more beneficial to improve lipid reserve and nutritional status than strictly decrease cholesterol levels for better clinical outcomes. It calls for causality validation and mechanism research, and further randomized controlled trials are urgently warranted to explore the roles of serum RC level and nutritional status in patients with ischemic stroke.

## Abbreviations

LDL-C: low-density lipoprotein cholesterol

RC: remnant cholesterol

VLDL: very low-density lipoprotein

IDL: intermediate-density lipoprotein

COUNT: Controlling Nutritional Status

mRS: modified Rankin Scale

TC: total cholesterol

TG: triglyceride

NIHSS: National Institutes of Health Stroke Scale

LAA: large-artery atherosclerosis

CE: cardioembolism

SAO: small-vessel occlusion

TOAST: Trial of ORG 10172 in Acute Stroke Treatment

HDL-C: high-density lipoprotein cholesterol

eGFR: estimated glomerular filtration rate

RCSs: restricted cubic splines.

RMB: Chinese Yuan Renminbi

OR: odds ratio.

CI: confidence interval.

## Supplementary Information

Supplementary data to this article can be found online.

## Data Availability

Data available within the article or its supplementary materials.

## Acknowledgments

We thank volunteer patients for their support and involvement in the study.

## Author contributions

All seven authors contributed to the conception of the study. HN, YW and DW drafted the manuscript and performed statistical analyses. NY, MC and XM contributed to preparing the tables and figures. HN and JZ obtained the research funding and revised the manuscript. All authors read and approved the final manuscript.

## Availability of data and materials

The data that support the findings of this study are available from the corresponding author upon reasonable request.

---

**Fig. S1.**
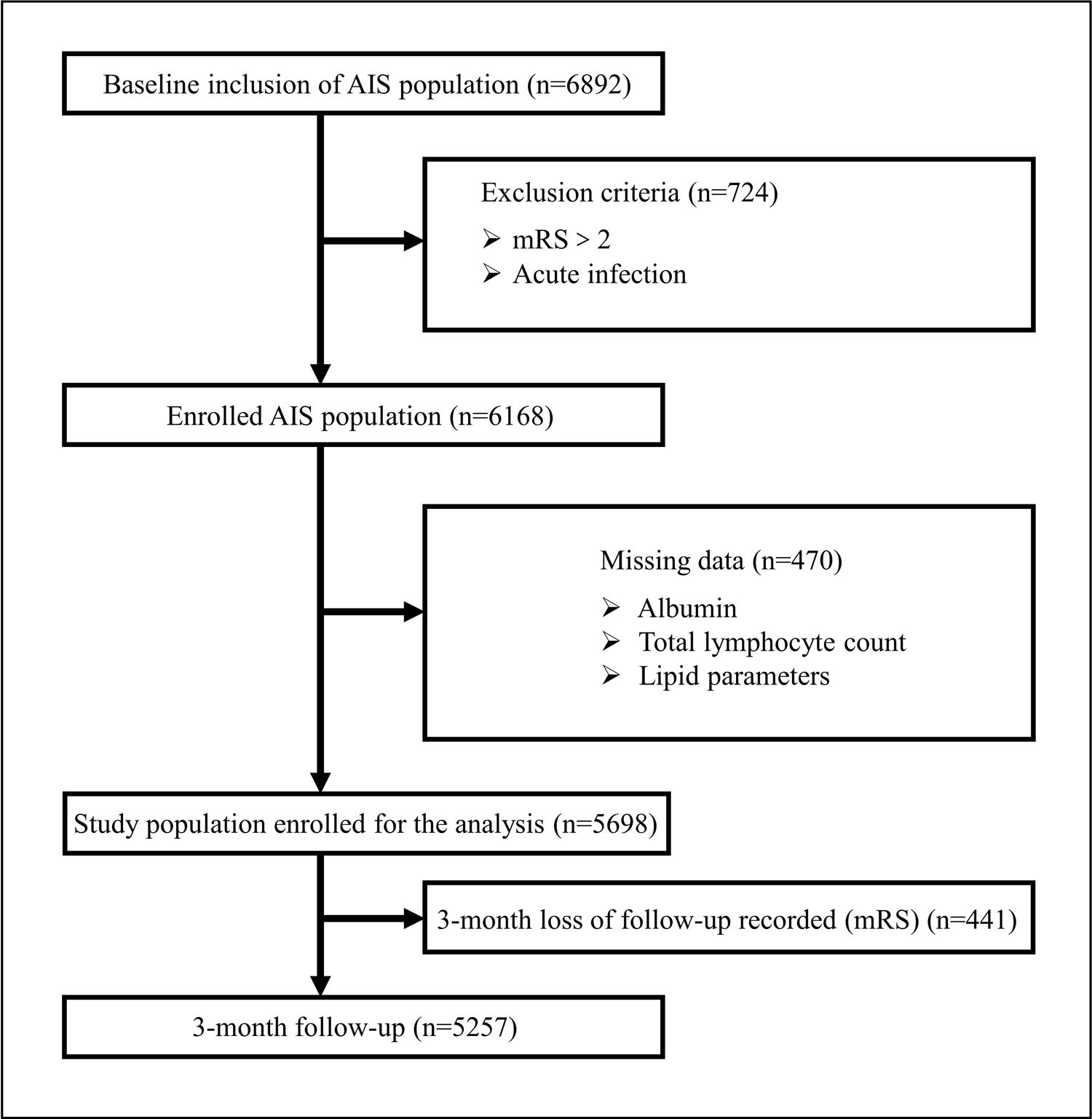
Flow chart of patient selection.

**Fig. S2.**
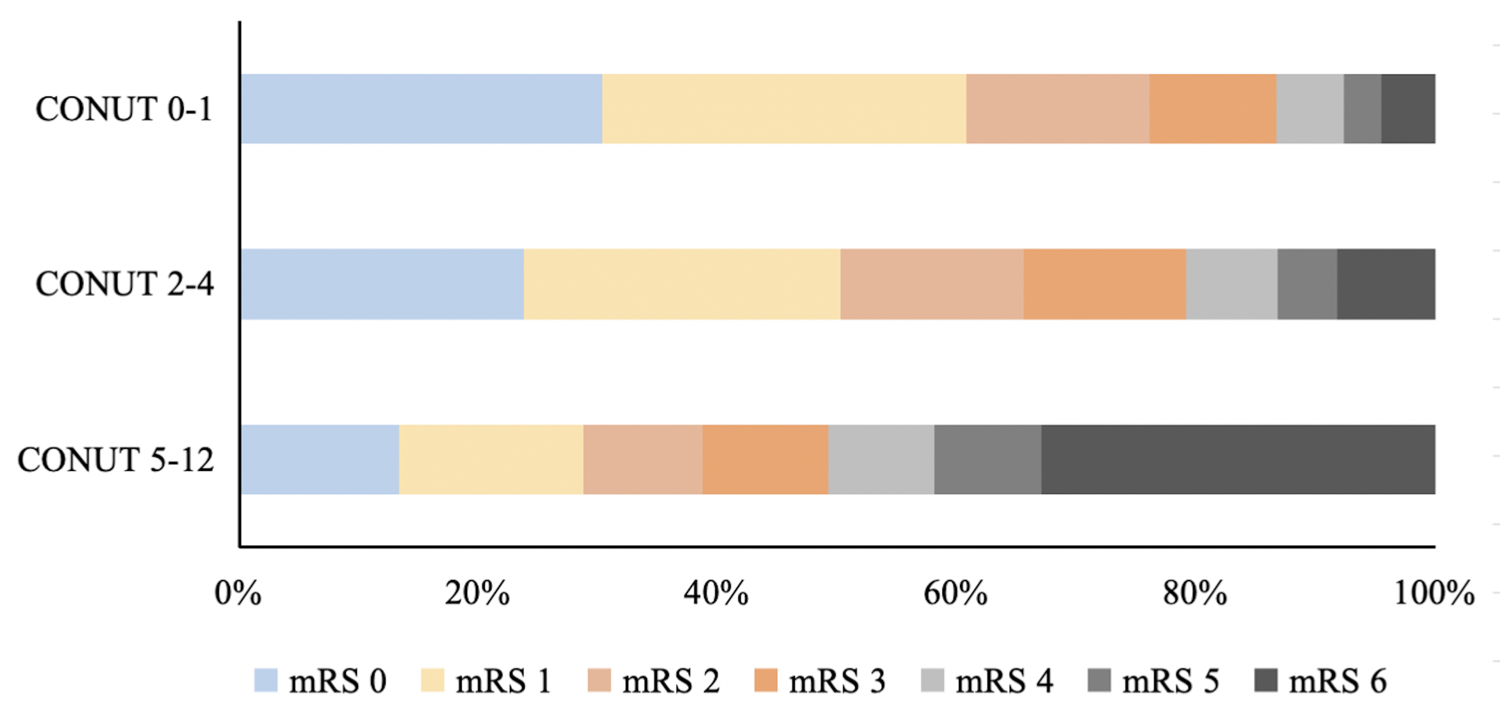
The distribution of stroke outcomes in different groups stratified by nutritional status. mRS, modified Rankin Scale.

**Fig. S3.**
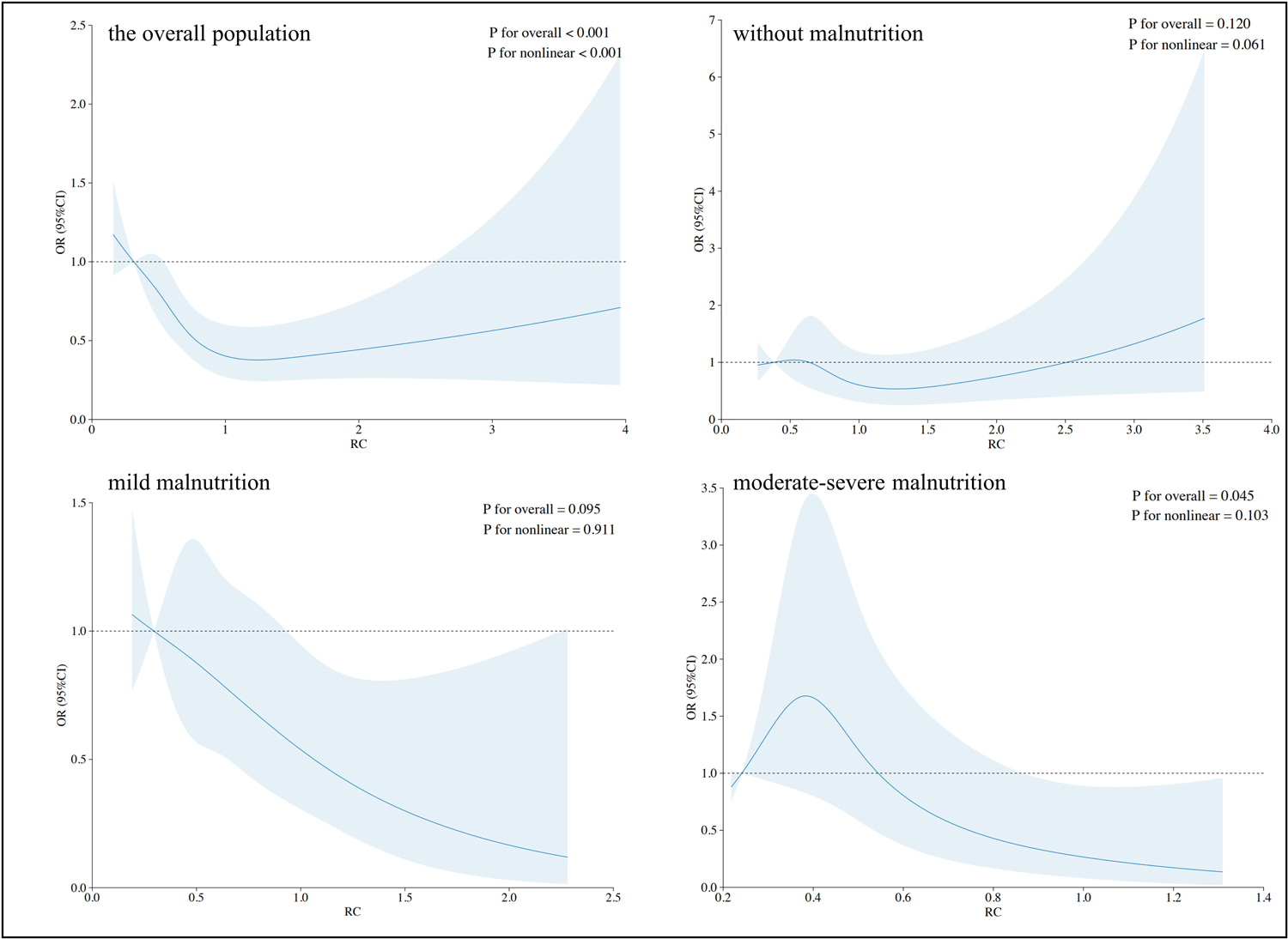
Distribution of RC level and its association between RC with all-cause mortality. Restricted cubic splines of RC with 4 predefined knots (5th, 35th, 65th, and 95th percentiles) to evaluate the non-linearity association of RC on a continuous scale and risk of all-cause mortality. These models were adjusted for age, gender, admission NIHSS scores, reperfusion therapy, smoking history, drinking history, hypertension, diabetes mellitus, prior stroke, hyperlipidemia, coronary artery disease, atrial fibrillation, stroke subtype, use of statin. RC, residual cholesterol.

**Fig. S4.**
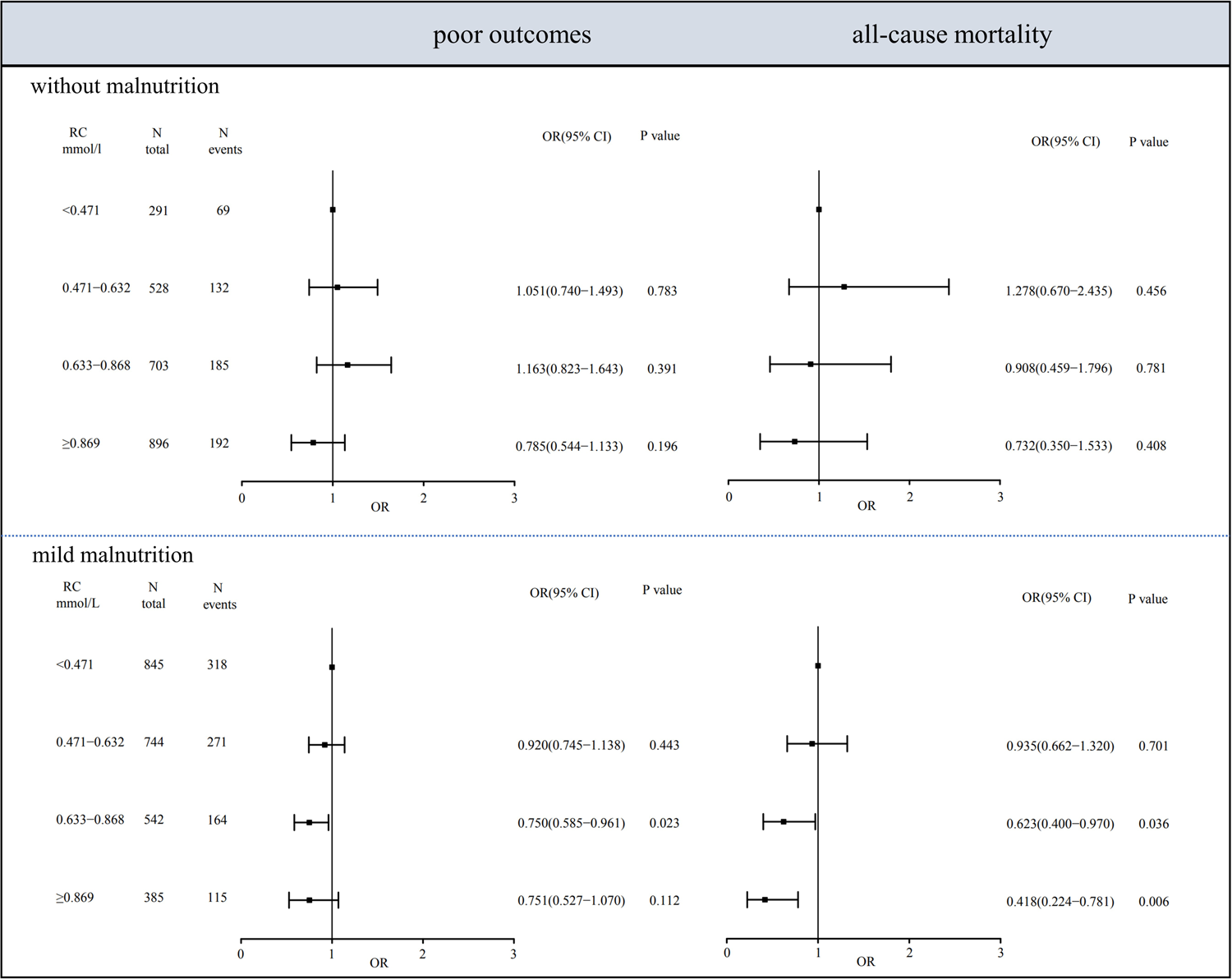
Association between RC in quartiles and risk of poor outcomes/all-cause mortality in patients without malnutrition and the mild malnourished. Adjusted for age, gender, admission NIHSS scores, reperfusion therapy, smoking history, drinking history, hypertension, diabetes mellitus, prior stroke, hyperlipidemia, coronary artery disease, atrial fibrillation, stroke subtype, use of statin. RC, residual cholesterol; OR, odds ratio; CI, confidence interval.

**Fig. S5.**
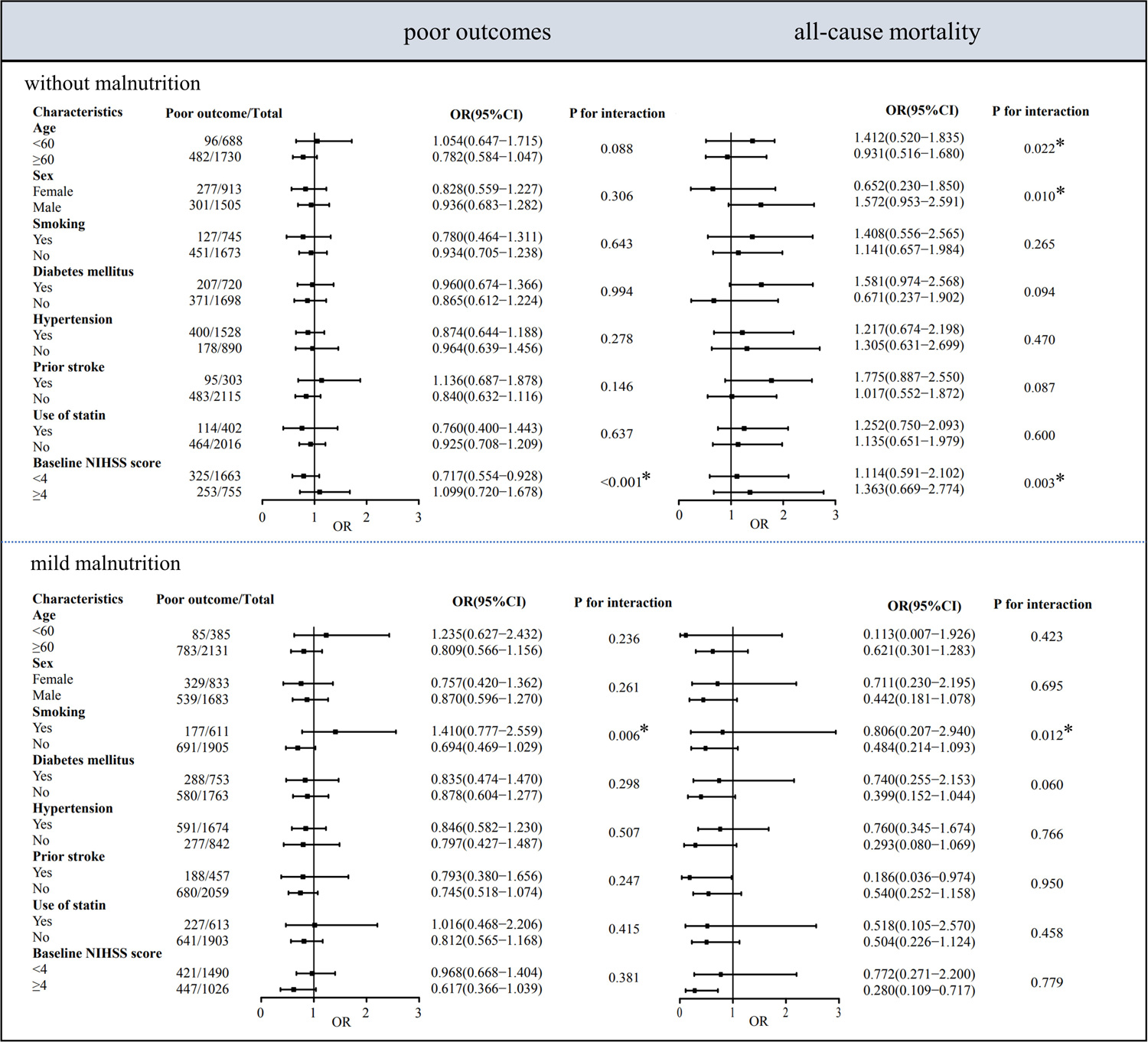
Subgroup analyses of the association between RC and poor outcomes/all-cause mortality in patients without malnutrition and the mild malnourished (quartile 4 vs quartile 1, data for quartiles 2 and 3 not shown). Adjusted for age, gender, admission NIHSS scores, reperfusion therapy, smoking history, drinking history, hypertension, diabetes mellitus, prior stroke, hyperlipidemia, coronary artery disease, atrial fibrillation, stroke subtype, use of statin, unless the variable was used as a subgroup variable, RC, residual cholesterol. OR, odds ratio; CI, confidence interval.

## References

1. Andone S, Farczádi L, Imre S and Bă laș a R. Fatty Acids and Lipid Paradox-Neuroprotective Biomarkers in Ischemic Stroke. International Journal of Molecular Sciences. 2022;23. doi: 10.3390/ijms231810810.

2. Langsted A, Madsen CM and Nordestgaard BG. Contribution of remnant cholesterol to cardiovascular risk. Journal of Internal Medicine. 2020;288:116–127. doi: 10.1111/joim.13059.

3. Li W, Huang Z, Fang W, Wang X, Cai Z, Chen G, Wu W, Chen Z, Wu S and Chen Y. Remnant Cholesterol Variability and Incident Ischemic Stroke in the General Population. Stroke. 2022;53:1934–1941. doi: 10.1161/strokeaha.121.037756.

4. Quispe R, Martin SS, Michos ED, Lamba I, Blumenthal RS, Saeed A, Lima J, Puri R, Nomura S, Tsai M, Wilkins J, Ballantyne CM, Nicholls S, Jones SR and Elshazly MB. Remnant cholesterol predicts cardiovascular disease beyond LDL and ApoB: a primary prevention study. European Heart Journal. 2021;42:4324–4332. doi: 10.1093/eurheartj/ehab432.

5. Tian Y, Wu Y, Qi M, Song L, Chen B, Wang C, Lu J, Yang Y, Zhang X, Cui J, Xu W, Yang H, He W, Zhang Y, Zheng X, Zhang H, Guo Y and Li X. Associations of remnant cholesterol with cardiovascular and cancer mortality in a nationwide cohort. Science Bulletin. 2024;69:526–534. doi: 10.1016/j.scib.2023.12.035.

6. Yang XH, Zhang BL, Cheng Y, Fu SK and Jin HM. Association of remnant cholesterol with risk of cardiovascular disease events, stroke, and mortality: A systemic review and meta-analysis. Atherosclerosis. 2023;371:21–31. doi: 10.1016/j.atherosclerosis.2023.03.012.

7. Fujihara Y, Nakamura T, Horikoshi T, Obata J-e, Fujioka D, Watanabe Y, Watanabe K and Kugiyama K. Remnant Lipoproteins Are Residual Risk Factor for Future Cardiovascular Events in Patients With Stable Coronary Artery Disease and On-Statin Low-Density Lipoprotein Cholesterol Levels <70 mg/dL. Circulation Journal. 2019;83:1302–1308. doi: 10.1253/circj.CJ-19-0047.

8. Varbo A and Nordestgaard BG. Remnant Cholesterol And Risk Of Ischemic Stroke In 112,512 Individuals From The General Population. Atherosclerosis. 2019;287:e85–e86. doi: 10.1016/j.atherosclerosis.2019.06.248.

9. Cheng K-H, Lin J-R, Anderson CS, Lai W-T and Lee T-H. Lipid Paradox in Statin-Naïve Acute Ischemic Stroke But Not Hemorrhagic Stroke. Frontiers in Neurology. 2018;9. doi: 10.3389/fneur.2018.00541.

10. Ho L-C, Wang H-K, Chiu L-T, Wang H-H, Lee Y-C, Hung S-Y, Sun Y, Wei C-Y, Hsu K-C, Chen Y-W, Lien L-M and Hsu CY. Protein energy wasting–based nutritional assessment predicts outcomes of acute ischemic stroke and solves the epidemiologic paradox. Nutrition. 2022;93. doi: 10.1016/j.nut.2021.111431.

11. Wang S-S, Yang S-S, Pan C-J, Wang J-H, Li H-W, Chen S-M, Hao J-K, Li X-H, Li R-R, Li B-Y, Yang J-H, Shi Y-T, Li H-H, Bao Y-H, Wang W-C, Du S-Y, He Y, Li C-L and Liu M. Cholesterol paradox in the community-living old adults: is higher better? Journal of Geriatric Cardiology. 2023;20:837–844. doi: 10.26599/1671-5411.2023.12.003.

12. Wang A, Zhang Y, Xia G, Tian X, Zuo Y, Chen P, Wang Y, Meng X and Han X. Association of serum albumin to globulin ratio with outcomes in acute ischemic stroke. Cns Neuroscience & Therapeutics. 2023;29:1357–1367. doi: 10.1111/cns.14108.

13. Adams HP, Bendixen BH, Kappelle LJ, Biller J, Love BB, Gordon DL and Marsh EE. Classification of subtype of acute ischemic stroke. Definitions for use in a multicenter clinical trial. TOAST. Trial of Org 10172 in Acute Stroke Treatment. Stroke. 1993;24:35–41. doi: 10.1161/01.str.24.1.35.

14. Friedewald WT, Levy RI and Fredrickson DS. Estimation of the concentration of low-density lipoprotein cholesterol in plasma, without use of the preparative ultracentrifuge. Clin Chem. 1972;18:499–502. doi: 10.1093/clinchem/18.6.499.

15. Wu W, Chen G, Wu K, Zheng H, Chen Y, Wang X, Huang Z, Cai Z, Cai Z, Chen Z, Lan Y, Chen S, Wu S and Chen Y. Cumulative exposure to high remnant-cholesterol concentrations increases the risk of cardiovascular disease in patients with hypertension: a prospective cohort study. Cardiovascular Diabetology. 2023;22. doi: 10.1186/s12933-023-01984-4.

16. Kong G, Zhang A, Chong B, Lim J, Kannan S, Chin YH, Ng CH, Lin C, Khoo CM, Muthiah M, Dalakoti M, Kristanto W, Wang Y, Kong W, Poh KK, Chai P, Foo R, Chan MY-Y, Loh P-H and Chew NWS. Long-Term Prognosis of Patients With Coexisting Obesity and Malnutrition After Acute Myocardial Infarction: A Cohort Study. Circulation-Cardiovascular Quality and Outcomes. 2023;16. doi: 10.1161/circoutcomes.122.009340.

17. Ding Q, Liu S, Yao Y, Liu H, Cai T and Han L. Global, Regional, and National Burden of Ischemic Stroke, 1990-2019. Neurology. 2022;98:E279–E290. doi: 10.1212/wnl.0000000000013115.

18. Sască u R, Clement A, Radu R, Prisacariu C and Stă tescu C. Triglyceride-Rich Lipoproteins and Their Remnants as Silent Promoters of Atherosclerotic Cardiovascular Disease and Other Metabolic Disorders: A Review. Nutrients. 2021;13. doi: 10.3390/nu13061774.

19. Nordestgaard BG and Varbo A. Triglycerides and cardiovascular disease. The Lancet. 2014;384:626–635. doi: 10.1016/s0140-6736(14)61177-6.

20. Duell PB. Triglyceride-Rich Lipoproteins and Atherosclerotic Cardiovascular Disease Risk. Journal of the American College of Cardiology. 2023;81:153–155. doi: 10.1016/j.jacc.2022.11.013.

21. Huh JH, Roh E, Lee SJ, Ihm S-H, Han K-D and Kang JG. Remnant Cholesterol Is an Independent Predictor of Type 2 Diabetes: A Nationwide Population-Based Cohort Study. Diabetes Care. 2023;46:305–312. doi: 10.2337/dc22-1550.

22. Duran EK and Pradhan AD. Triglyceride-Rich Lipoprotein Remnants and Cardiovascular Disease. Clinical Chemistry. 2021;67:183–196. doi: 10.1093/clinchem/hvaa296.

23. Navarese EP, Vine D, Proctor S, Grzelakowska K, Berti S, Kubica J and Raggi P. Independent Causal Effect of Remnant Cholesterol on Atherosclerotic Cardiovascular Outcomes: A Mendelian Randomization Study. *Arteriosclerosis*, Thrombosis, and Vascular Biology. 2023;43. doi: 10.1161/atvbaha.123.319297.

24. Bhatt DL, Steg PG, Miller M, Brinton EA, Jacobson TA, Ketchum SB, Doyle RT, Juliano RA, Jiao L, Granowitz C, Tardif J-C and Ballantyne CM. Cardiovascular Risk Reduction with Icosapent Ethyl for Hypertriglyceridemia. New England Journal of Medicine. 2019;380:11–22. doi: 10.1056/NEJMoa1812792.

25. Baigent C, Blackwell L, Emberson J, Holland LE, Reith C, Bhala N, Peto R, Barnes EH, Keech A, Simes J, Collins R and Cholesterol Treatment T. Efficacy and safety of more intensive lowering of LDL cholesterol: a meta-analysis of data from 170 000 participants in 26 randomised trials. Lancet. 2010;376:1670–1681. doi: 10.1016/s0140-6736(10)61350-5.

26. Zuliani G, Cherubini A, Atti AR, Ble A, Vavalle C, Di Todaro F, Benedetti C, Volpato S, Marinescu MG, Senin U and Fellin R. Low cholesterol levels are associated with short-term mortality in older patients with ischemic stroke. *The journals of gerontology Series A*, Biological sciences and medical sciences. 2004;59:293–7. doi: 10.1093/gerona/59.3.m293.

27. Chen L, Xu J, Sun H, Wu H and Zhang J. The total cholesterol to high-density lipoprotein cholesterol as a predictor of poor outcomes in a Chinese population with acute ischaemic stroke. Journal of Clinical Laboratory Analysis. 2017;31. doi: 10.1002/jcla.22139.

28. Ha SH, Ryu J-C, Bae J-H, Koo S, Kwon B, Song Y, Lee DH, Chang JY, Kang D-W, Kwon SU, Kim JS and Kim BJ. High serum total cholesterol associated with good outcome in endovascular thrombectomy for acute large artery occlusion. Neurological Sciences. 2022;43:5985–5991. doi: 10.1007/s10072-022-06269-4.

29. Vauthey C, de Freitas GR, van Melle G, Devuyst G and Bogousslavsky J. Better outcome after stroke with higher serum cholesterol levels. Neurology. 2000;54:1944–9. doi: 10.1212/wnl.54.10.1944.

30. Tuttolomondo A, Di Raimondo D, Di Sciacca R, Pedone C, La Placa S, Arnao V, Pinto A and Licata G. Effects of clinical and laboratory variables at admission and of in-hospital treatment with cardiovascular drugs on short term prognosis of ischemic stroke. The GIFA study. Nutrition Metabolism and Cardiovascular Diseases. 2013;23:642–649. doi: 10.1016/j.numecd.2012.01.010.

31. Olsen TS, Christensen RHB, Kammersgaard LP and Andersen KK. Higher total serum cholesterol levels are associated with less severe strokes and lower all-cause mortality - Ten-year follow-up of ischemic strokes in the Copenhagen Stroke Study. Stroke. 2007;38:2646–2651. doi: 10.1161/strokeaha.107.490292.

32. Satou R, Matsuzawa Y, Akiyama E, Konishi M, Yoshii T, Okada K, Maejima N, Iwahashi N, Hibi K, Kosuge M, Ebina T, Tamura K and Kimura K. Low-density lipoprotein cholesterol levels on admission and long-term outcomes in statin-naive patients with acute coronary syndrome. European Heart Journal. 2020;41:1647–1647. doi: 10.1093/ehjci/ehaa946.1647.

33. Sato R, Matsuzawa Y, Yoshii T, Akiyama E, Konishi M, Nakahashi H, Minamimoto Y, Kimura Y, Okada K, Maejima N, Iwahashi N, Kosuge M, Ebina T, Kimura K, Tamura K and Hibi K. Impact of Low-Density Lipoprotein Cholesterol Levels at Acute Coronary Syndrome Admission on Long-Term Clinical Outcomes. Journal of Atherosclerosis and Thrombosis. 2024;31:444–460. doi: 10.5551/jat.64368.

34. Niu H, Chu M, Yang N, Wang D, Liu Y, Mao X, Xia S, Wang D, Wu X and Zhao J. Prognosis of patients with coexisting obesity and malnutrition after ischemic stroke: A cohort study. Clinical Nutrition. 2024;43:1171–1179. doi: 10.1016/j.clnu.2024.04.005.

35. Davis CJ, Sowa D, Keim KS, Kinnare K and Peterson S. The Use of Prealbumin and C-Reactive Protein for Monitoring Nutrition Support in Adult Patients Receiving Enteral Nutrition in an Urban Medical Center. Journal of Parenteral and Enteral Nutrition. 2012;36:197–204. doi: 10.1177/0148607111413896.

36. Ishida S, Hashimoto I, Seike T, Abe Y, Nakaya Y and Nakanishi H. Serum albumin levels correlate with inflammation rather than nutrition supply in burns patients: a retrospective study. The journal of medical investigation: JMI. 2014;61:361–8. doi: 10.2152/jmi.61.361.

37. Mukai H, Villafuerte H, Qureshi AR, Lindholm B and Stenvinkel P. Serum albumin, inflammation, and nutrition in end-stage renal disease: C-reactive protein is needed for optimal assessment. Seminars in Dialysis. 2018;31:435–439. doi: 10.1111/sdi.12731.

38. Yuan K, Zhu S, Wang H, Chen J, Zhang X, Xu P, Xie Y, Zhu X, Zhu W, Sun W, Xu G and Liu X. Association between malnutrition and long-term mortality in older adults with ischemic stroke. Clinical Nutrition. 2021;40:2535–2542. doi: 10.1016/j.clnu.2021.04.018.

39. Zhang G, Pan Y, Zhang R, Wang M, Meng X, Li Z, Li H, Wang Y, Zhao X, Liu G and Wang Y. Prevalence and Prognostic Significance of Malnutrition Risk in Patients With Acute Ischemic Stroke: Results From the Third China National Stroke Registry. Stroke. 2022;53:111–119. doi: 10.1161/strokeaha.121.034366.

40. Chen Y, Yang H, Lan M, Wei H and Chen Y. The controlling nutritional status score and risk factors associated with malnutrition in patients with acute ischemic stroke. Frontiers in Neurology. 2023;14. doi: 10.3389/fneur.2023.1067706.

41. Lu H-Y, Ho U-C and Kuo L-T. Impact of Nutritional Status on Outcomes of Stroke Survivors: A Post Hoc Analysis of the NHANES. Nutrients. 2023;15. doi: 10.3390/nu15020294.

42. Lu Y-W, Lu S-F, Chou R-H, Wu P-S, Ku Y-C, Kuo C-S, Chang C-C, Tsai Y-L, Wu C-H and Huang P-H. Lipid paradox in patients with acute myocardial infarction: Potential impact of malnutrition. Clinical Nutrition. 2019;38:2311–2318. doi: 10.1016/j.clnu.2018.10.008.

43. Zhou L, Zhang H, Wang S, Zhao H, Li Y, Han J, Zhang H, Li X and Qu Z. PCSK-9 inhibitors: a new direction for the future treatment of ischemic stroke. Frontiers in Pharmacology. 2024;14. doi: 10.3389/fphar.2023.1327185.

44. Amarenco P, Kim JS, Labreuche J, Charles H, Giroud M, Lee B-C, Lavallee PC, Mahagne M-H, Meseguer E, Nighoghossian N, Steg PG, Vicaut E, Bruckert E and Treat Stroke Target I. Yield of Dual Therapy With Statin and Ezetimibe in the Treat Stroke to Target Trial. Stroke. 2022;53:3260–3267. doi: 10.1161/strokeaha.122.039728.

